# A Machine Learning Approach to Defining and Predicting the Scale of Typhoid Fever Outbreaks

**DOI:** 10.64898/2026.03.19.26348781

**Authors:** Zeaan Pithawala, Jo Walker, Dae-Hyup Koh, Jong-Hoon Kim, Virginia E. Pitzer

## Abstract

Despite improvements in access to clean water and sanitation, typhoid fever outbreaks continue to cause substantial morbidity and mortality worldwide. The World Health Organization (WHO) recommends the use of typhoid conjugate vaccines (TCVs) for outbreak response; however, guidance on when to trigger such a response remains unclear due to the absence of a standardized definition of a typhoid fever outbreak. We analyzed a dataset of 34 typhoid fever outbreaks with detailed time series data between 2000 and 2022 and found that all documented outbreaks lasted at least 7 days and were associated with at least 6 total cases or at least 2 laboratory-confirmed cases. Applying unsupervised machine learning methods to this dataset revealed two distinct clusters: one containing relatively large outbreaks (≥288 total cases), and the other containing relatively small outbreaks (≤191 total cases). We then labeled 215 typhoid outbreaks from the same period as either large or small using a simple threshold of 250 total cases and trained supervised machine learning models to predict the outbreak size classification on the basis of country-level features in the year before the outbreak. We found that large outbreaks are most likely to occur in countries where access to safely managed sanitation services, the proportion of urban dwellers, and basic drinking water services are low. We then used these models to project the magnitude of typhoid outbreaks for 192 countries in 2023. We identified several high-risk areas in Africa and South Asia that were consistently predicted to face large-scale outbreaks across most of the models. These findings provide evidence-based criteria for defining the onset of an outbreak, enabling the triggering of timely responses to typhoid outbreaks and preparation for potential large-scale epidemics.

## Introduction

Typhoid fever outbreaks continue to occur worldwide from multiple sources, such as contaminated food and water, and have also resulted from the emergence of antimicrobial-resistant strains [1, 2, 3]. Meanwhile, typhoid fever remains endemic in many low- and middle-income countries (LMICs) and is estimated to cause 10 to 20 million cases and over 100,000 deaths annually [4, 5], mostly in Africa and Asia.

The World Health Organization (WHO) has recommended the use of typhoid conjugate vaccines (TCVs) in countries with a high burden of typhoid fever and/or high prevalence of antimicrobial resistance as well as in response to confirmed outbreaks of typhoid fever [6, 7]. In addition, Gavi has provided funding support for the introduction of TCVs in high-burden countries, resulting in the vaccination of over 100 million children to date [8, 9]. However, data to guide the use of TCV in response to outbreaks are very limited. Efforts are needed to assess the value of both preventive and reactive vaccination campaigns for outbreaks of typhoid fever.

A previous study that explored the potential health impact and cost-effectiveness of deploying TCVs in response to a typhoid fever outbreak found that the reactive deployment of TCVs during an outbreak could be a cost-effective approach provided that the delays in vaccine deployment are minimal [10]. However, there is no definitive threshold for triggering timely TCV vaccination due to limited public health surveillance of typhoid [10, 11]. Thus, defining the start of a typhoid fever outbreak is crucial for achieving optimal public health outcomes.

Machine learning offers a promising approach for detecting and predicting the size of typhoid fever outbreaks. While machine learning has been applied to tasks such as screening asymptomatic travelers for SARS-CoV-2 at border checkpoints [12], forecasting influenza trends [13], and classifying viral genomes [14], to our knowledge, no study has examined its use in identifying common patterns among typhoid outbreaks or in predicting their potential scale. Applying machine learning methods to identify countries susceptible to large-scale typhoid outbreaks and establishing a threshold to trigger a timely response could therefore support country-level preparedness and response to typhoid outbreaks.

In this analysis, we developed a set of criteria for defining typhoid fever outbreaks by applying unsupervised machine learning methods to a comprehensive dataset of typhoid outbreaks with time series data from 2000 to 2022. We also trained supervised machine learning models to predict the size of typhoid fever outbreaks given baseline country-level data on water, sanitation, and hygiene (WaSH) coverage, demographics, and air travel patterns.

## Methods

### Typhoid outbreak data

We used a comprehensive dataset of typhoid fever outbreaks that occurred from 2000 to 2022 with reported time series data to identify criteria for defining and classifying outbreaks of typhoid fever [15]. This dataset comprises 34 distinct outbreaks across 21 countries and includes metadata such as case counts (suspected, probable, and confirmed cases), start and end dates of the outbreak, dates of any public health interventions, peak case dates, and geographic information at the city, country, and WHO region levels. To eliminate redundancy, we removed duplicate outbreak entries, defined as identical outbreaks reported across different studies or the same outbreaks reported at multiple temporal resolutions (daily, weekly, and monthly). In the case of the latter, we always retained the outbreak record with the greatest temporal resolution (daily case counts if available, or weekly). Additional outbreak-level variables were derived using the available date fields in the dataset. Specifically, we calculated *outbreak duration* as the number of days between the start and end dates, *time to peak* as the number of days between the start and peak dates, and *time to intervention* as the number of days between the start and reported intervention dates.

To maintain data integrity and minimize potential bias from imputation, we eliminated variables for which over 20% of outbreaks had a missing value, including *attack rate*, and the number of *total hospitalizations*, *total complications*, and *total deaths*. Of the remaining variables, missing values of *total suspected cases* (∼9% missing data, n = 3), *total confirmed cases* (∼9% missing data, n = 3), and *time to peak* (∼3% missing data, n = 1) were imputed using iterative imputation with a Random Forest regressor (100 estimators, max 10 iterations). This method is robust to non-linear interactions and preserves the internal correlation structure of the data. Categorical variables, such as *WHO region*, were encoded using the one-hot method.

To reduce multicollinearity and ensure more reliable clustering outcomes, we excluded the *total suspected cases* and *time to peak* variables from the clustering analysis because they displayed high collinearity (r > 0.8) with *total cases* (r = 0.97) and *outbreak duration* (r = 0.94), respectively. We excluded *total confirmed cases* from the clustering analysis to avoid biases arising from heterogeneity in case reporting across countries. We also excluded the *AMR status* and *Country* variables from the clustering analysis, as our objective is to define a typhoid outbreak based on case burden and temporal characteristics rather than features tied to specific antimicrobial resistance profiles or country. Thus, the final set of variables included *total cases*, *outbreak duration*, and *WHO region*, allowing us to characterize outbreaks based on their magnitude, duration, and location.

### Clustering

An overview of the analysis pipeline is provided in Figure 1. After cleaning and processing the dataset, we used unsupervised machine learning to identify clusters of outbreaks with distinct characteristics. Our first step was to identify the optimal number of distinct clusters in the dataset. Selecting too many clusters (overclustering) could separate similar outbreaks into different groups, making it difficult to recognize common features. Conversely, selecting too few clusters (underclustering) could cause dissimilar outbreaks to be grouped together, reducing our ability to identify real differences. We evaluated a range from 1 to 34 clusters, with the former representing a single cluster containing all outbreaks, and the latter representing a scenario in which each outbreak forms its own cluster. For each of these 34 scenarios, we performed dimensionality reduction using Uniform Manifold Approximation and Projection (UMAP) [16] and classified outbreaks into clusters using the K-means algorithm, which minimizes within-cluster variance by iteratively assigning observations to the nearest cluster centroid and recalculating the centroids until convergence is achieved. We then used two approaches to evaluate the support for each possible number of clusters: the silhouette score method and the elbow method.

**Figure 1:**
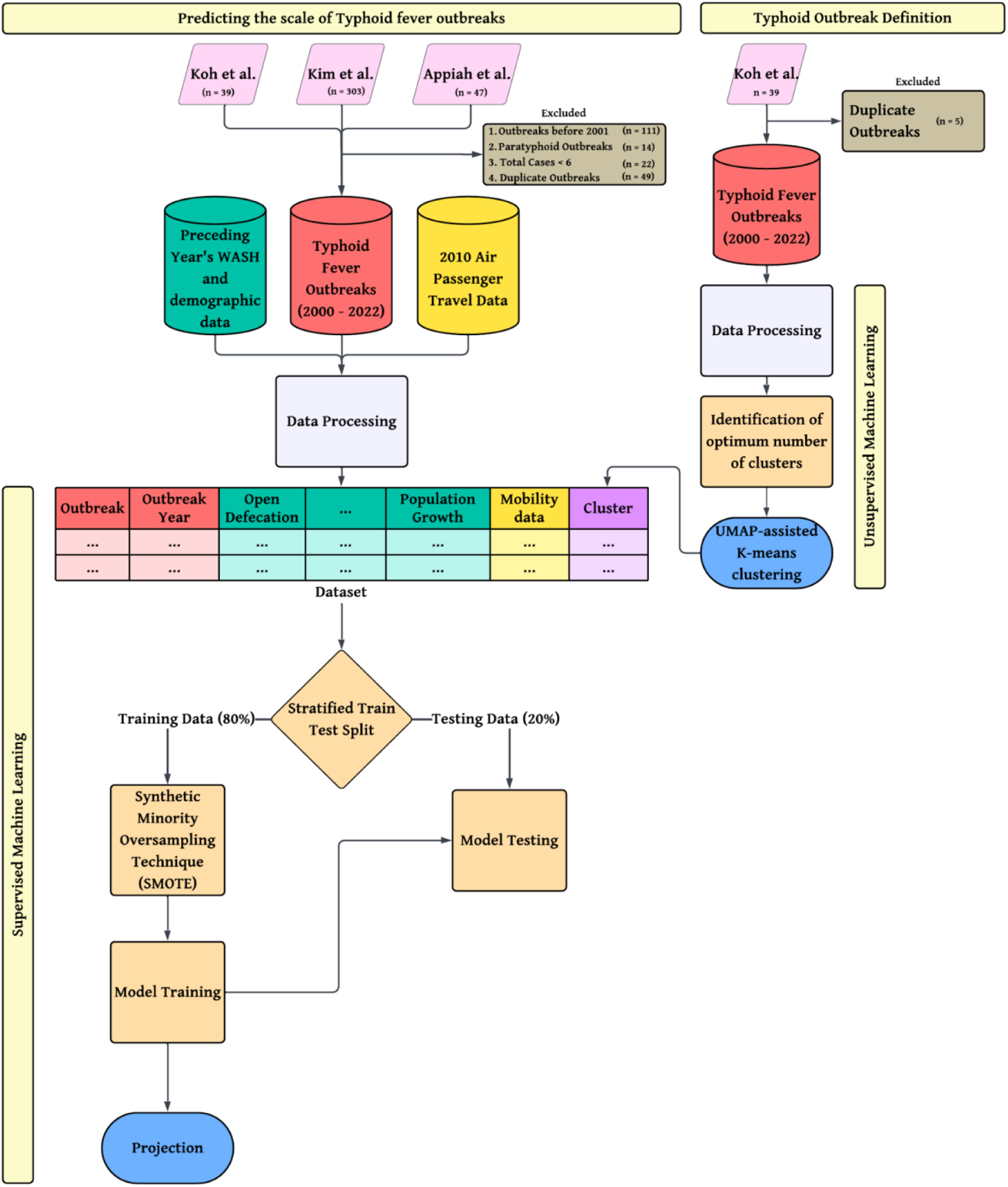
Pipeline of the analysis. Unsupervised machine learning was used to group outbreaks from the Koh et al. dataset into clusters. We then used the derived clustering rule to label a larger set of outbreaks and trained supervised machine learning models to predict the scale of outbreaks given baseline WaSH, demographic, and air travel data.

A silhouette score quantitatively measures how well each data point fits within its assigned cluster compared to other clusters. For an individual outbreak *i*, the silhouette score (*Si*) is defined as:

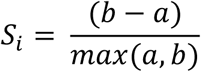

where *a* is the mean Euclidean distance between the *i*^th^ outbreak and all other outbreaks in the same cluster, and *b* is the mean Euclidean distance between the *i*^th^ outbreak and all the outbreaks of the next closest cluster. Silhouette scores range from −1 to +1, where a negative score indicates that the outbreak is closer to another cluster than its own cluster, a score of 0 means that the outbreak is equally close to its own cluster and the closest other cluster, and a positive score indicates that the outbreak is closer to the center of its own cluster and is far away from other clusters. Using the silhouette score method, we defined the optimal number of clusters as that which maximizes the mean silhouette score across all outbreaks.

As an additional indicator of the appropriate number of clusters, we also used the elbow method, which evaluates the total within-cluster sum of squares (WCSS, also known as inertia) for varying numbers of clusters. As the number of clusters increases, the inertia naturally decreases. However, beyond a certain point, the rate of improvement sharply declines, creating an “elbow” in the curve. By visually inspecting the plot of inertia versus the number of clusters, we identified the inflection point, which suggests an optimal trade-off between model complexity and explained variance. Both methods consistently pointed toward two meaningful clusters, which informed our subsequent clustering.

Following the identification of the optimal number of clusters, we used a UMAP-assisted K-means clustering approach to classify each of the 34 outbreaks into the most appropriate cluster. We applied UMAP over Principal Component Analysis (PCA) for dimensionality reduction due to its ability to preserve both local and global structures in the data and to better capture potential non-linear relationships within the high-dimensional feature space. UMAP projects the data into two dimensions, facilitating visualization and reducing noise that could skew meaningful patterns.

To assess the robustness of the UMAP-assisted K-means clustering approach, we performed the clustering procedure 1,000 times using different random seeds to account for the stochastic nature of UMAP embeddings. For each iteration, the dataset was embedded using UMAP and clustered using K-means with *k*=2. We then computed a consensus clustering by identifying, for each sample, the most frequent cluster assignment across all 10,000 runs. Remarkably, this consensus solution exactly matched the original clustering result. This high level of consistency indicates that the observed clusters are not artifacts of random variation introduced by UMAP’s stochastic optimization, but rather reflect a stable and reproducible structure in the data, which strongly supports the validity and reproducibility of the identified clusters.

To evaluate the quality of the final cluster classification, we calculated the mean silhouette score, the Davies-Bouldin index, and the Calinski-Harabasz index. These metrics assess the compactness and separation of clusters. We calculated descriptive statistics to characterize the size, timing, and location of outbreaks in the two fitted clusters.

### Predicting the scale of typhoid fever outbreaks

We merged the Koh et al. dataset with two additional datasets (Kim et al. [17] and Appiah et al. [1]), which describe global typhoid fever outbreaks but do not include time series data. We excluded outbreaks that took place before 2001 due to limited WaSH data from this period, paratyphoid outbreaks, outbreaks involving fewer than six reported cases (the smallest outbreak in Koh et al.), and duplicate outbreak entries, retaining the record with time series data if available. After excluding non-qualifying outbreaks, we had 215 outbreaks in our dataset. We labeled each outbreak in the merged dataset as either large (≥250 total reported cases) or small (<250 total reported cases). The threshold of 250 cases was based on the clustering results described above.

We linked each outbreak to country-level WaSH and demographic data from the World Bank for the year preceding the outbreak. We used country-level data because consistent subnational-level data were not available for all outbreaks, and using country-level indicators allowed us to train models that are generalizable across countries. These variables included: *proportion of people using at least basic drinking water services*, *proportion of people using safely managed drinking water sources*, *proportion of people using at least basic sanitation services*, *proportion of people using safely managed sanitation services*, *prevalence of open defecation*, *proportion of people with basic handwashing facilities*, *population growth*, *proportion of people living in urban areas* and *proportion of people living in rural areas*. These variables were reported for the total population and further disaggregated into urban and rural populations. We also included a country-level variable representing the proportion of incoming international air travelers from high typhoid-incidence countries. We calculated this variable using modeled air travel data [18] and country-level typhoid incidence estimates [19]. This air travel data was only available for the year 2010; therefore, we assumed the relative volume of air travel between countries is fairly stable over time.

As a categorical variable, we encoded the *WHO region* using the one-hot method. Variables with more than 20% missing data were excluded from the analysis. For variables with less than 20% missing data, missing values were imputed using an iterative imputation method. As described above, we excluded highly collinear features based on a predefined threshold (Pearson correlation > 0.8) to minimize multicollinearity in the dataset.

The dataset was split into training (80%) and testing (20%) sets using stratified sampling to maintain class distribution. To address class imbalance, Synthetic Minority Oversampling Technique (SMOTE) [20] was applied to the training data only, generating synthetic samples for the minority class while preserving the original test set for unbiased evaluation. We trained Logistic Regression, k-Nearest Neighbors (kNN), Support Vector Machine (SVM), Random Forest, and XGBoost models to the data (Table 1).

**Table 1:** Model parameters.

| Machine Learning Classifier | Parameter Specification |
| --- | --- |
| Logistic Regression | {max_iter=1000, random_state=100} |
| k-Nearest Neighbour (kNN) | {n_neighbors=5} |
| Support Vector Machine (SVM) | {kernel='rbf', C=1.0, probability=True, random_state=100} |
| Random Forest | {n_estimators=1000, random_state=100} |
| XGBoost | {n_estimators=1000, use_label_encoder=False, eval_metric='logloss', random_state=100} |

Each model outputs the predicted probability that a given outbreak is “large” (versus “small”). By default, a classification threshold of 0.5 is used to convert these predicted probabilities into binary classifications (that is, outbreaks with predicted probability > 0.5 are classified as large, and those with predicted probability ≤ 0.5 as small). Rather than using the default classification probability threshold of 0.5 to classify outbreaks as “small” or “large”, we determined the optimal classification probability threshold for each model using Youden’s J statistic (sensitivity + specificity – 1) [21]. This approach systematically evaluates all possible thresholds and selects the one that maximizes the combined performance of sensitivity and specificity, which is particularly important when dealing with imbalanced datasets. Using these optimized thresholds, we computed key performance metrics, including accuracy, recall, precision, F1-score, and Matthews Correlation Coefficient (MCC).

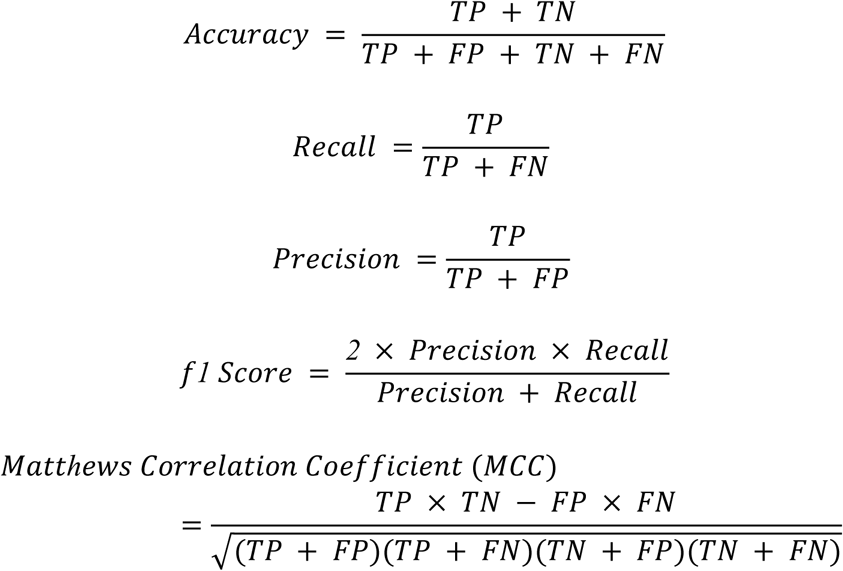

where TP is True Positives, FP is False Positives, TN is True Negatives, and FN is False Negatives.

To provide robust performance estimates, five-fold stratified cross-validation was conducted, and SMOTE was applied to each fold’s training dataset to maintain consistency with the training procedure and avoid distributional mismatch between model training and cross-validation. This approach was chosen due to the imbalanced nature of the dataset and the absence of group-related dependencies. Mean area under the curve (AUC) and accuracy scores were computed with 95% confidence intervals, enabling reliable model comparison while maintaining class balance within each fold.

The trained models were applied to a dataset of 192 countries with variables based on 2022 data, to predict the scale of typhoid fever outbreaks in 2023, followed by an out-of-sample model validation.

Finally, we conducted sensitivity analyses to assess the robustness of our findings to our definition of a large outbreak (≥ 250 cases). We evaluated two alternative thresholds to classify large outbreaks: ≥ 100 cases and ≥ 500 cases. In each scenario, outbreaks were reclassified according to the specified cutoff, and the analyses were repeated accordingly.

### Explainability of machine learning models

Machine learning models are often considered black boxes, making it difficult to assess the influence of individual variables on predictions. To enhance interpretability and better understand the contribution of covariates, we applied SHapley Additive exPlanations (SHAP) [22] to the XGBoost and Random Forest models, which were the two most complex and least interpretable among the five machine learning algorithms evaluated. We focused on countries in the Oceania region, where greater variability in predictions was observed, and examined the influence of covariates on outbreak scale classifications in these settings.

## Results

### Typhoid outbreak definition

Among the 34 unique outbreaks identified in the Koh et al. dataset, each involved a minimum of six total cases or two blood-culture-confirmed cases, and all the outbreaks lasted at least seven days.

Both the silhouette score and elbow methods indicated that two clusters is the optimal number. This was the number of clusters which maximized the silhouette score and for which there was an inflection point in inertia (Figure 2).

**Figure 2:**
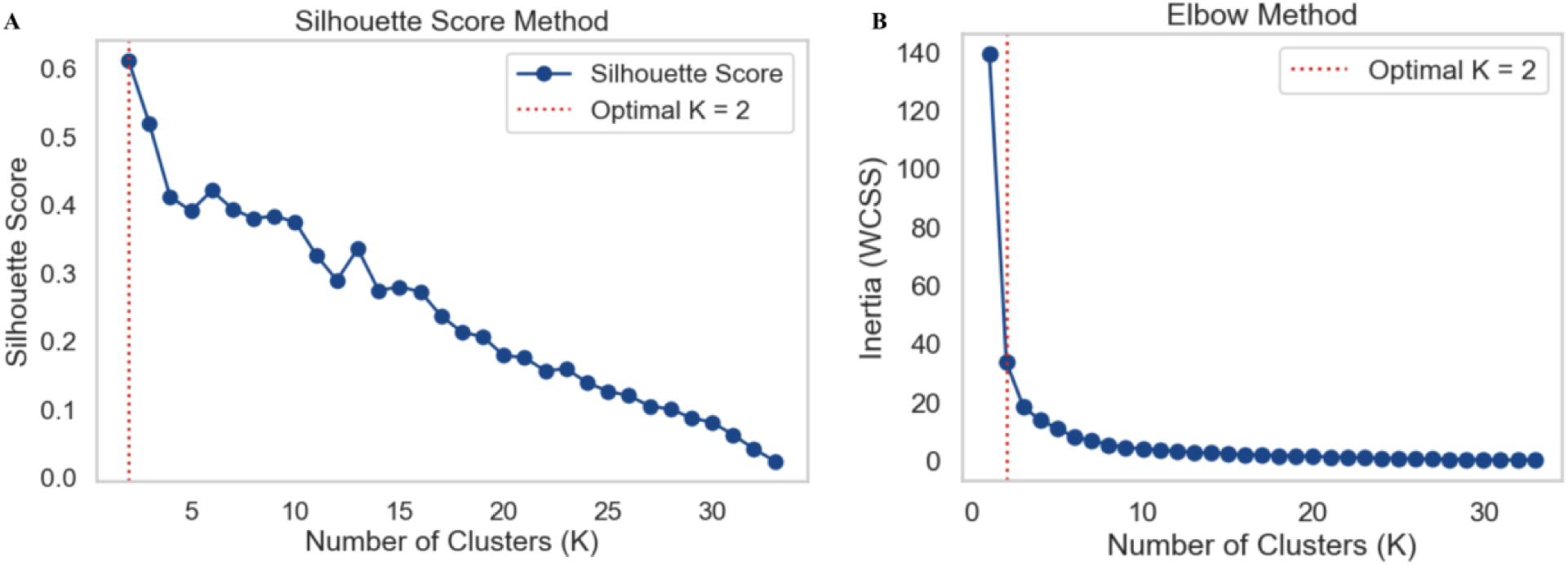
Identification of the Optimal Number of Outbreak Clusters using the Silhouette Score and Elbow Methods. | Prior to clustering the data using unsupervised machine learning methods, we determined the optimal number of clusters (k) using two approaches: (A) the silhouette score method and (B) the elbow method. Despite their methodological differences, both approaches converged on the same optimal number of clusters (k=2). Note that the silhouette score is undefined when there is only a single cluster. WCSS: within-cluster sum of squares.

After determining that two outbreak clusters were optimal, we applied UMAP-assisted K-means clustering with k=2 clusters, which resulted in two well-balanced clusters of typhoid fever outbreaks. Cluster *A* contained 19 outbreaks, while Cluster *B* included 15 outbreaks (Figure 3).

**Figure 3:**
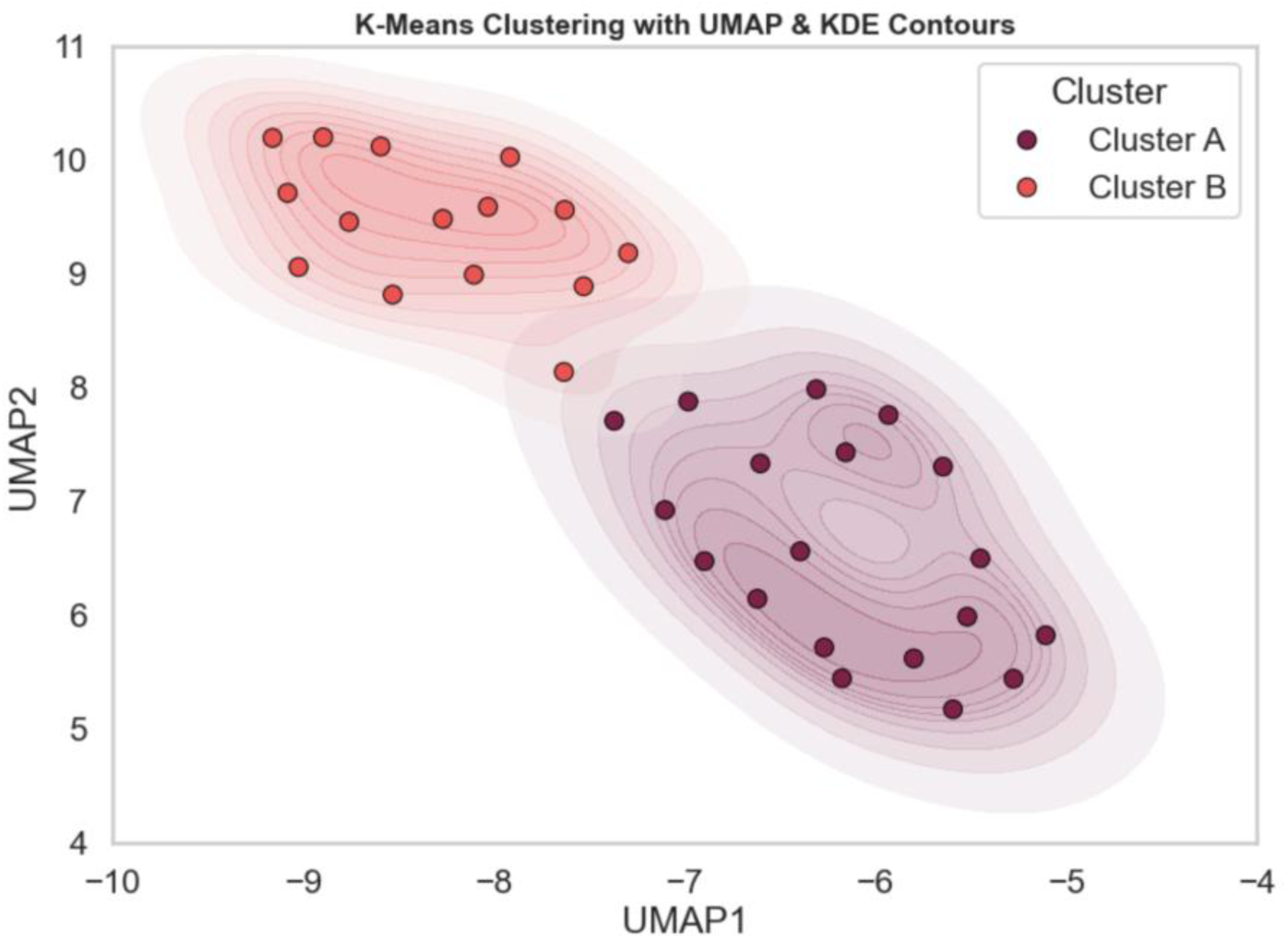
UMAP-assisted K-means Clustering of Typhoid Fever Outbreaks from 2000 - 2022 with Reported Time Series Data. | The clustering results show a clear separation into two distinct clusters, suggesting the presence of two underlying types of typhoid fever outbreak patterns. This distinction can be attributed to the total number of typhoid cases in these outbreaks. Outbreaks in Cluster *A* (or “Large outbreak cluster”) had at least 288 typhoid cases (which includes suspected, probable and confirmed cases), while outbreaks in Cluster *B* (or “Small outbreak cluster”) had at most 191 cases.

Clustering performance was evaluated using the silhouette score, Davis-Bouldin index, and Calinski-Harabasz index. The silhouette score was 0.61, indicating good separation between the clusters. The Davis-Bouldin and Calinski-Harabasz index values were 0.51 and 100.89, respectively, further supporting the validity of the clustering structure (for these non-negative indices, lower values indicate better clustering performance).

Outbreaks in cluster A contained a median of 879 total cases (range: 288 to 10,152) and lasted for a median of 181 days (range: 14 to 1,460) (Table 2). The outbreaks in cluster B were smaller and shorter in comparison, containing a median of 75 cases (range: 6 to 191) with a median duration of 60 days (range: 7 to 311).

**Table 2:**
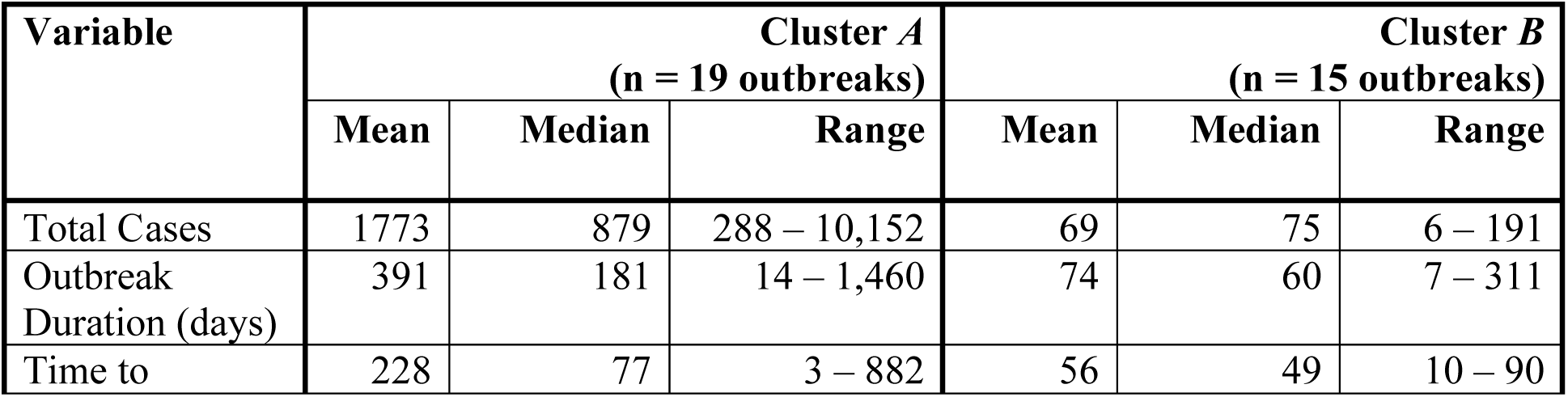
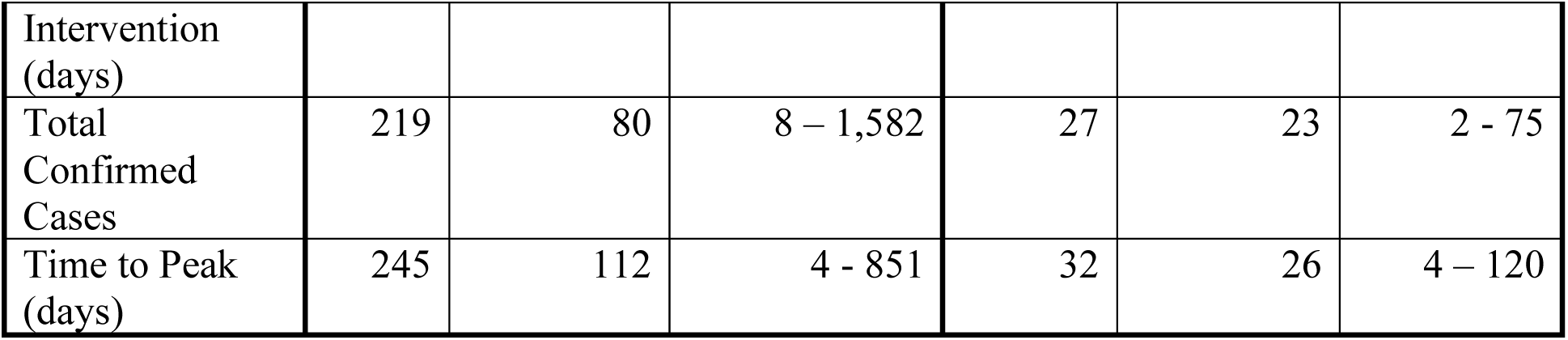
Characteristics of each cluster within the Koh et al. dataset.

The highest number of cases observed in the “small outbreak” cluster was 191. Recognizing that smaller outbreaks are less likely to be reported due to their limited public health impact and frequent under-detection, we incorporated a 30% allowable margin of error to account for potential underreporting. Therefore, we defined outbreaks as “small” if the total number of cases (including suspected, probable, and laboratory-confirmed cases) was fewer than 250 and “large” if the total number exceeded 250.

### Predicting the scale of typhoid fever outbreaks

After excluding variables that had too many missing observations (>20% of outbreaks) or were highly collinear (*r* > 0.8), the final set of predictive features included *WHO region*, *proportion of international travelers arriving from high typhoid incidence countries*, *proportion of people using safely managed sanitation services*, *proportion of people using at least basic drinking water services*, *prevalence of open defecation*, *total population growth*, and the *share of the population living in urban areas*.

Three features contained missing values - *proportion of international travelers arriving from high typhoid incidence countries* (5.2% of outbreaks), *proportion of people using safely managed sanitation services* (17.6% of outbreaks), and *prevalence of open defecation* (0.5% of outbreaks) - which were imputed using iterative imputation with a multivariate approach. Distributions before and after imputation showed no substantial deviation (Figure S1).

Prior to balancing the dataset using SMOTE, the training set consisted of 116 small outbreaks (∼75%) and 41 large outbreaks (∼25%), indicating class imbalance. SMOTE generated a balanced training dataset with 116 examples per class. The testing set consisted of 30 small outbreaks and 10 large outbreaks.

Across all models, XGBoost, an ensemble method, gave the highest overall accuracy, with kNN following closely. Logistic regression attained the highest F1-score and MCC score. Overall, all the models had comparable results during model training (Table 3).

**Table 3:** Outbreak Classification Performance on Training Data.

| Machine Learning | Optimal Classification | Accuracy | Precision | Recall | F1-score | MCC | AUC |
| --- | --- | --- | --- | --- | --- | --- | --- |

| Classifier | Probability Threshold |  |  |  |  |  |  |
| --- | --- | --- | --- | --- | --- | --- | --- |
| Logistic Regression | 0.31 | 0.56 | 0.36 | 0.90 | 0.51 | 0.32 | 0.64 |
| k-Nearest Neighbour | 0.60 | 0.64 | 0.38 | 0.60 | 0.46 | 0.23 | 0.66 |
| Support Vector Machine | 0.46 | 0.59 | 0.33 | 0.60 | 0.43 | 0.16 | 0.58 |
| Random Forest | 0.18 | 0.56 | 0.35 | 0.80 | 0.48 | 0.25 | 0.63 |
| XGBoost | 0.04 | 0.67 | 0.40 | 0.60 | 0.48 | 0.26 | 0.62 |

The performance of five classification models was evaluated using stratified 5-fold cross-validation (Table 4). Among all models, SVM achieved the highest accuracy at 0.72 (95% CI: 0.63, 0.81), followed by kNN at 0.66 (95% CI: 0.59, 0.72). However, logistic regression yielded the highest AUC at 0.72 (95% CI: 0.68, 0.76), indicating the best overall discriminatory performance. Random Forest and SVM showed comparable AUCs at 0.71 (95% CI: 0.68, 0.74) and 0.71 (95% CI: 0.63, 0.80), respectively, despite Random Forest having lower accuracy (0.57 vs 0.72).

**Table 4:**
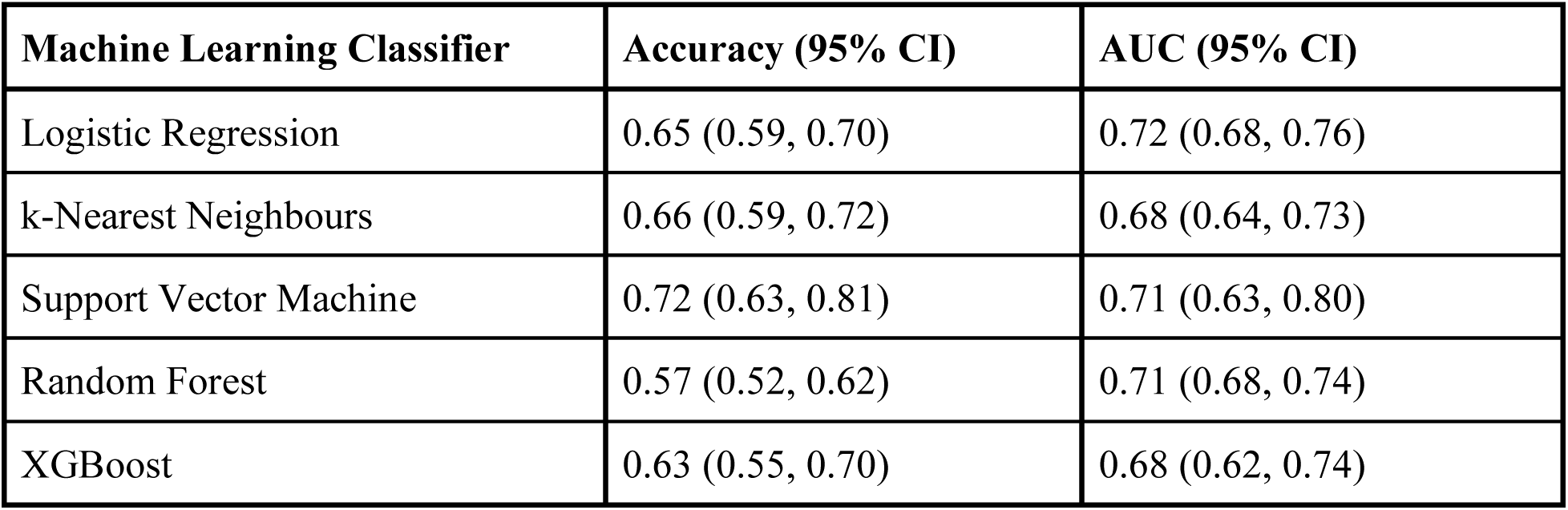
Classification Performance Under Stratified 5-Fold Cross Validation.

Each of the five model algorithms (Random Forest, XGBoost, Logistic Regression, SVM, kNN) demonstrated moderate to high performance in classifying typhoid fever outbreaks as “large” versus “small” during model training. Moreover, the classification thresholds of these models varied substantially. For instance, the optimal classification threshold for XGBoost was 0.039, while for kNN it was 0.600. This highlights the need to calibrate model-specific thresholds for classification, rather than relying on the default threshold of 0.5 (Figure 5). The models generally performed well in predicting large typhoid fever outbreaks (i.e., showed high sensitivity), minimizing false negatives.

Despite differences in their underlying algorithms, the models produced largely consistent predictions across most countries. In South Asian countries such as India, Bangladesh, Pakistan, and Nepal, as well as African nations including Malawi, the Democratic Republic of Congo, and Nigeria, the majority of the models predicted that, if a typhoid outbreak were to occur, it would likely be large in scale. Conversely, for countries in North America and most of Europe, the models consistently predicted a low likelihood of large-scale typhoid outbreaks (Figures 4, S2A, S2B).

**Figure 4:**
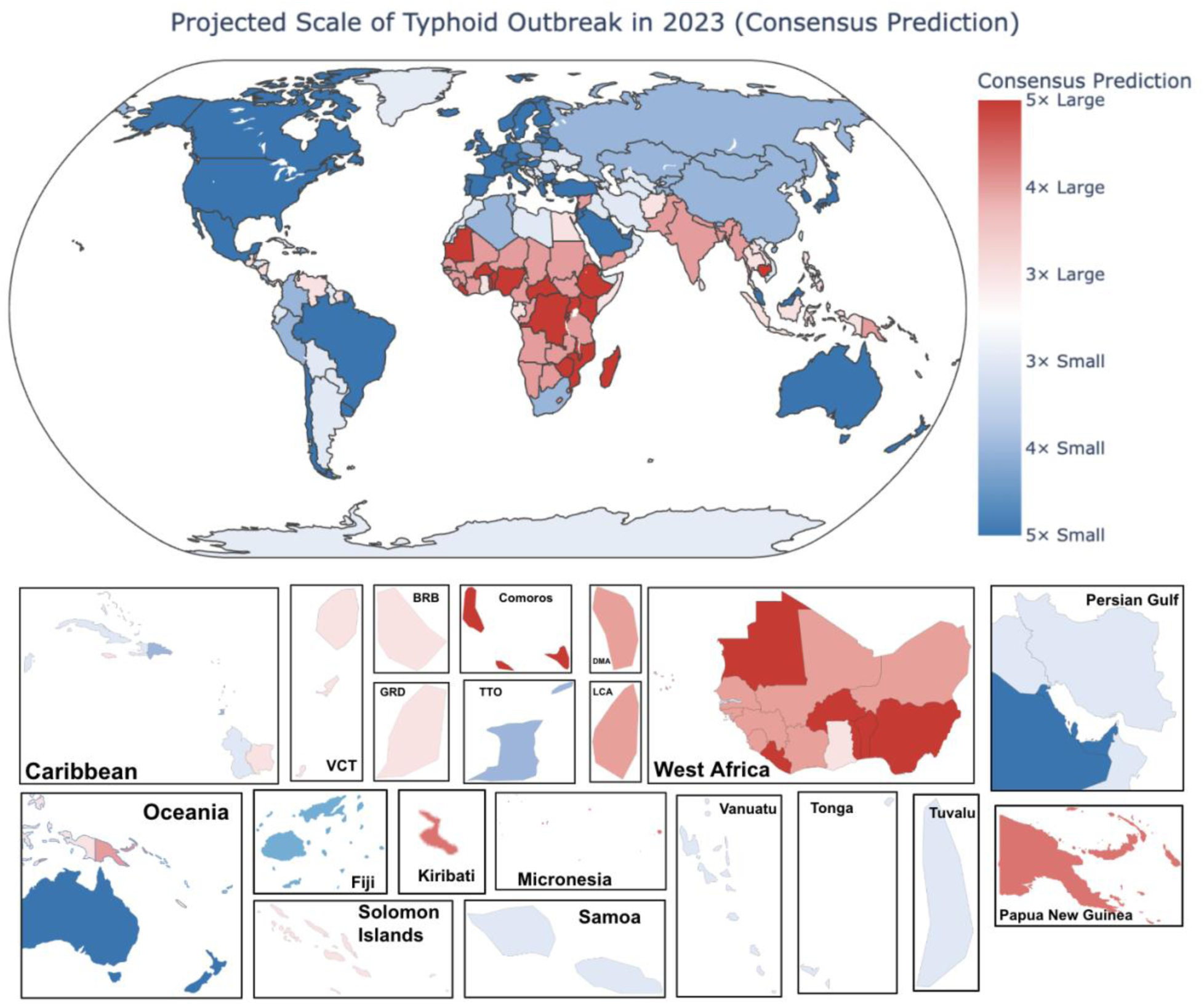
Consensus prediction of the scale of potential typhoid fever outbreaks in 2023. | Model agreement across five classifiers (Random forest, XGBoost, Support Vector Machine, logistic regression, and k-Nearest Neighbors), where each country is shaded according to the number of models predicting either a large outbreak (red) or a small outbreak (blue). Darker shades indicate stronger agreement among models, while lighter shades indicate greater uncertainty.

Figures 4, S2A, and S2B show the projected scale of typhoid fever outbreaks in 2023 by country. Typhoid outbreaks were consistently predicted to be large in sub-Saharan Africa, South Asia, and parts of South America, while outbreaks in North America, Europe, and the Western Pacific were usually predicted to be small. Disagreements amongst models in the scale of typhoid fever outbreaks were observed in Oceania and countries such as Guyana and Argentina in South America, and Mongolia in Central Asia. In Oceania, models were largely in agreement for Fiji, Australia, and New Zealand, suggesting less intense typhoid fever outbreaks; however, for Kiribati and Micronesia, the models consistently predicted large-scale outbreaks.

Figure 5 provides a more granular representation of model outputs. Higher probabilities (closer to 1, indicated with darker red) indicate stronger model confidence in the occurrence of a large-scale typhoid fever outbreak, whereas lower probabilities (closer to 0, indicated by darker blue) suggest that the model is confident in the occurrence of a small-scale typhoid fever outbreak.

**Figure 5:**
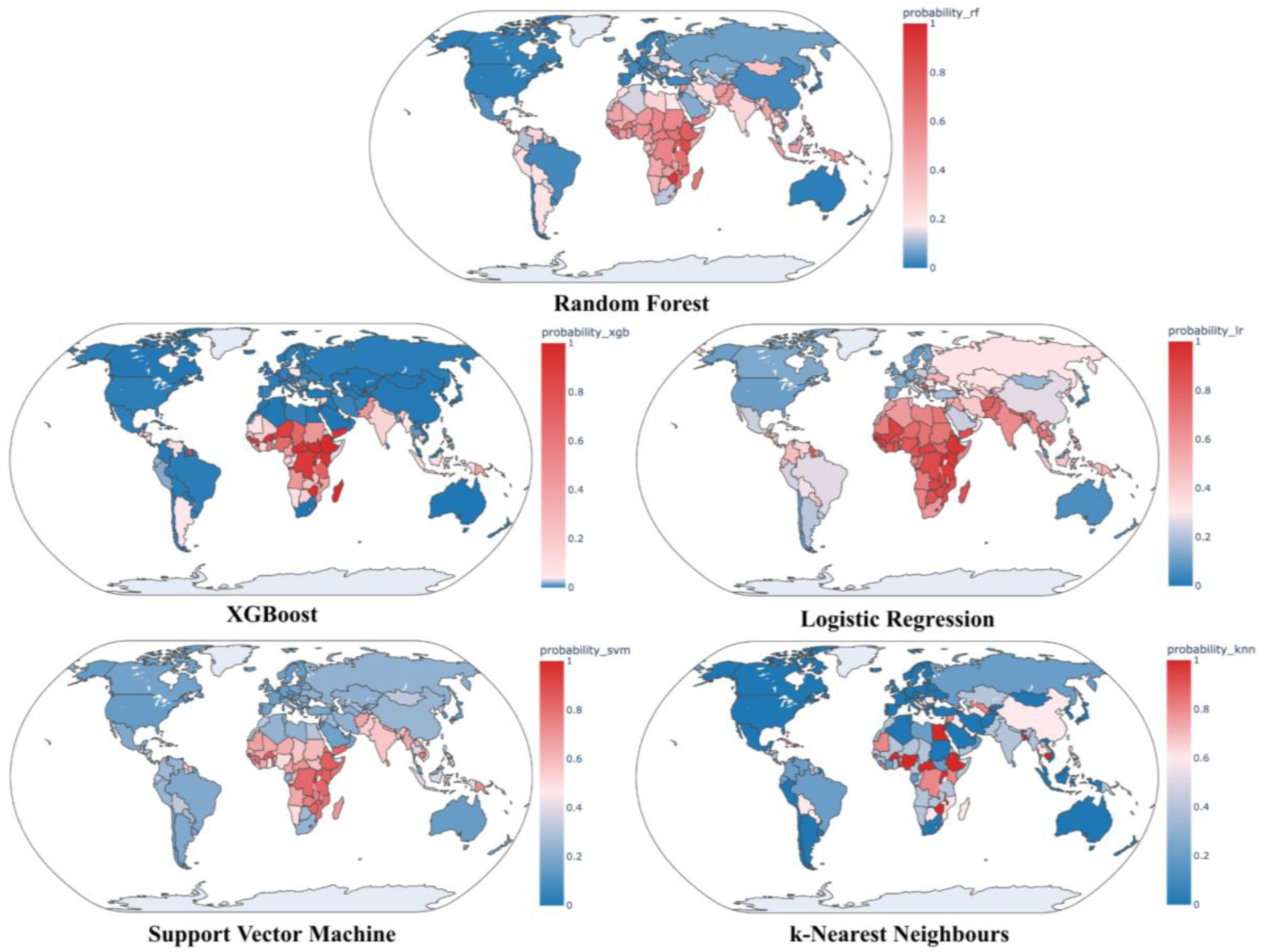
Predicted probabilities of potential large-scale typhoid fever outbreaks across the world in 2023. | Each map shows the predicted probabilities of countries into large outbreaks (red) or small outbreaks (blue) generated by five supervised learning algorithms: Random Forest, XGBoost, Logistic Regression, Support Vector Machine, and k-Nearest Neighbours. Instead of using the common classification threshold of 0.5 for each model, we calculated an optimal threshold for each model to balance sensitivity and specificity. Tree-based models such as XGBoost and Random Forest tended to have lower thresholds compared to distance-based models such as SVM and kNN. Despite these differences, the models performed similarly during cross-validation, and we observed that they were generally more confident in predicting large outbreaks in Africa.

Although the classification threshold varies by model, regions in South Asia and parts of sub-Saharan Africa consistently exhibited high predicted probabilities for large typhoid fever outbreaks. In contrast, regions such as North America, Europe, Australia, and New Zealand demonstrated low probabilities, corresponding to a low predicted outbreak scale. Regions in the Middle East and Central Asia showed probabilities closer to the classification thresholds (indicated by pink), reflecting greater uncertainty in model predictions for these areas.

Across all countries examined, Rwanda and Burundi were consistently ranked among the top ten with the highest probability of experiencing large typhoid outbreaks across all five models. In addition, we identified countries vulnerable to large outbreaks by WHO region. In the South-East Asia Region (SEAR), all five models consistently predicted high probabilities of large-scale outbreaks in Bangladesh, Nepal, India, Myanmar, Sri Lanka, Thailand, and Timor-Leste. In the African Region (AFR), Ethiopia, along with Rwanda and Burundi, was consistently highlighted. In the Eastern Mediterranean Region (EMR), Pakistan, Somalia, and Yemen were predicted to have the highest probabilities, while in the Western Pacific Region (WPR), Cambodia, Papua New Guinea, and the Federated States of Micronesia were consistently identified (Figure S3). The following countries were consistently predicted by all models to experience large typhoid fever outbreaks (Figure 4): Democratic Republic of Congo, Nigeria, Malawi, Zimbabwe, Uganda, Togo, Rwanda, Mozambique, Mauritius, Mauritania, Madagascar, Liberia, Kenya, Ethiopia, Comoros, Central African Republic, Cambodia, Burundi, Burkina Faso, and Benin. These findings suggest that establishing regional stockpiles of TCVs that can be quickly deployed in these countries may be critical to ensure preparedness and timely response to future typhoid fever outbreaks.

We identified five typhoid fever outbreaks reported in 2023, 2024, and 2025. Two were large-scale outbreaks, occurring in the Democratic Republic of Congo [23] and the Philippines [24], and three were small-scale outbreaks in New Zealand [25], Brazil [26], and Philippines [27]. Our model correctly predicted the scale of typhoid fever outbreaks in most of these settings (Table 5).

**Table 5:**
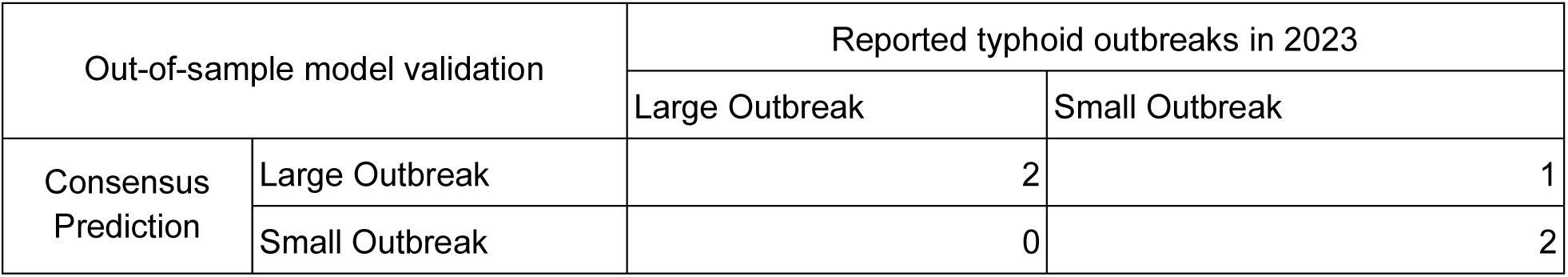
Out-of-Sample Validation of the Consensus Typhoid Outbreak Prediction Model (2023 - 2025)

Sensitivity analyses using alternative thresholds to define a large outbreak (≥100 and ≥500 cases) demonstrated that the overall geographic patterns were broadly consistent with the primary analysis (≥250 cases) (Figure 6).

**Figure 6:**
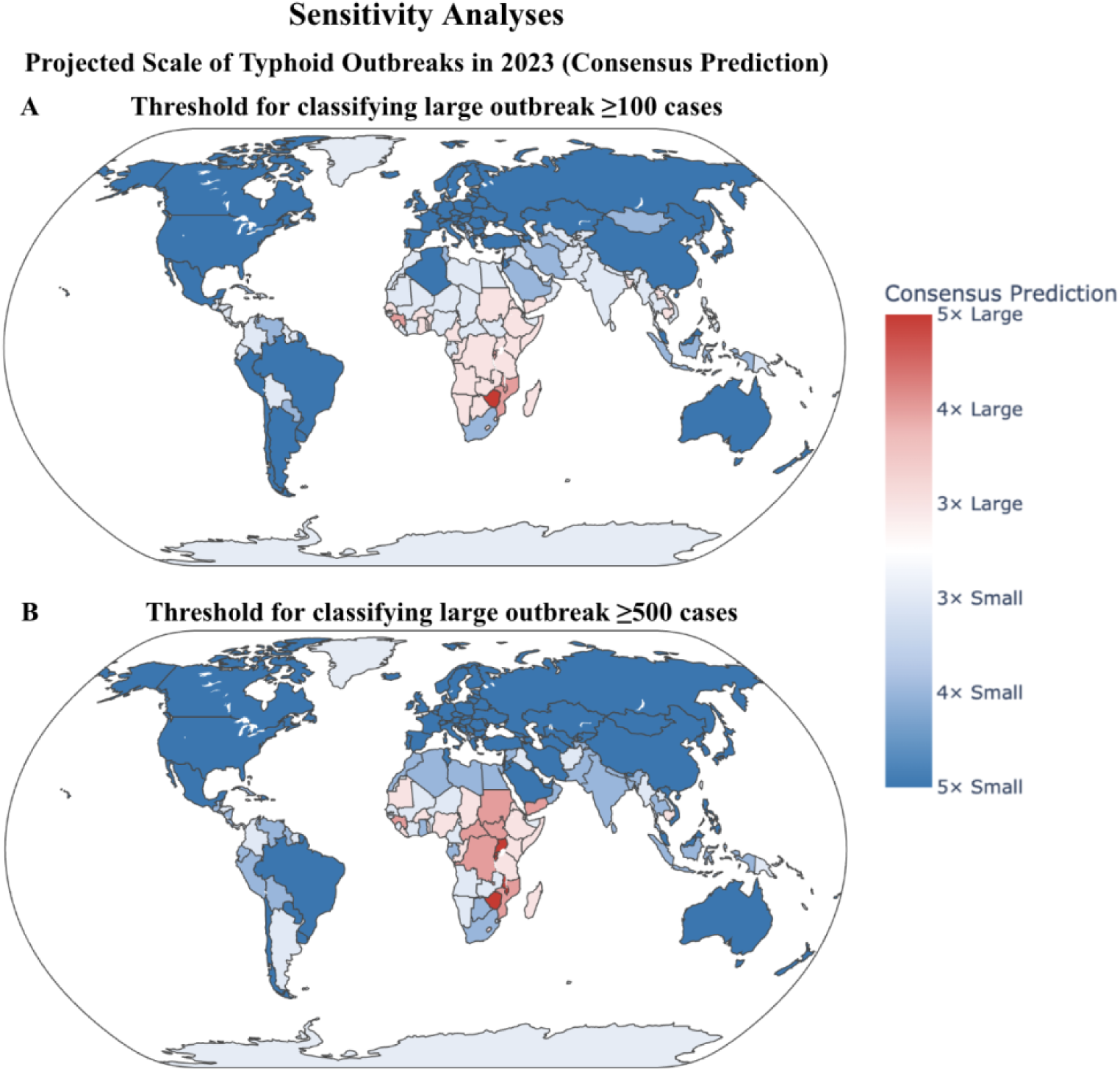
Sensitivity Analyses. Consensus predictions of the projected scale of potential typhoid fever outbreaks in 2023 under alternative definitions of a “large” outbreak: (A) ≥100 cases and (B) ≥500 cases. Each country is shaded according to the number of models predicting either a large outbreak (red) or a small outbreak (blue). Darker shades indicate stronger agreement among models, while lighter shades indicate greater uncertainty.

Across all thresholds, high-income countries were consistently predicted to have a low likelihood of large-scale typhoid outbreaks, whereas multiple countries in sub-Saharan Africa were predicted to be at high risk of large-scale outbreaks should an outbreak occur.

However, the classification of South Asian and Oceania countries varied by threshold. At the lower threshold (≥100 cases), fewer South Asian countries were identified as high risk compared to the primary scenario, and there were greater disagreements among models regarding the classification of countries in South Asia and Oceania compared to the higher threshold scenario (≥500 cases). Overall, we observed greater variability and less agreement among the models in the alternative scenarios compared with the primary scenario (Figures 4, 6, S4, and S5).

### Explainability of machine learning models

To evaluate the contribution of individual predictors to the model’s prediction, we applied SHAP to Random Forest (Figure 7A) and XGBoost (Figure 7B) models. These plots rank features by their importance and illustrate the directionality of their influence on the predicted outcome.

**Figure 7:**
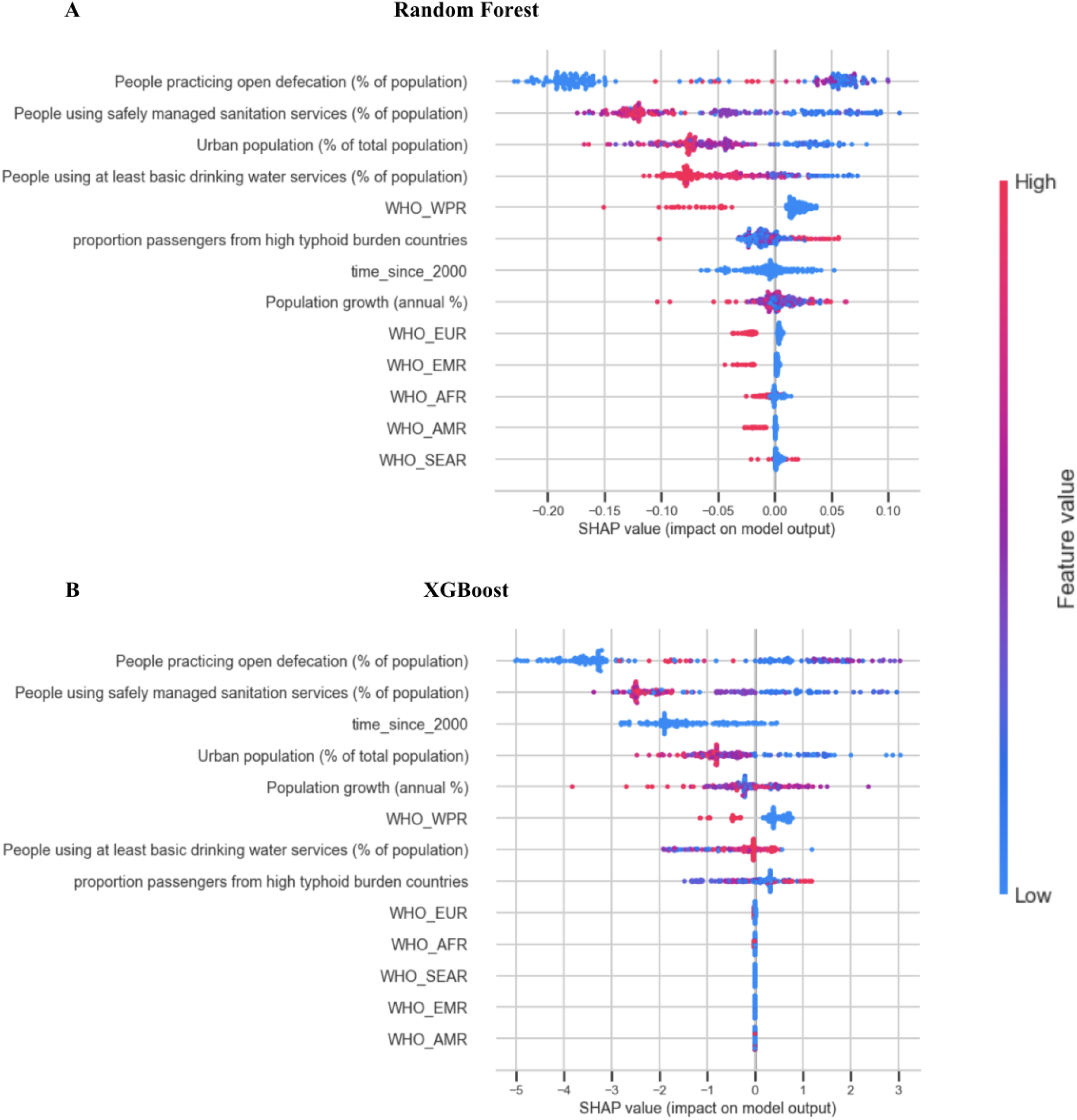
SHAP Summary Plots for Model Interpretability. SHAP summary plots for the (A) Random Forest and (B) XGBoost models, illustrating the contribution of covariates to predicted model outcomes. Features are ranked in descending order of global importance. Points to the right of the vertical zero-line represent an increase in the predicted probability of large outbreak, while points to the left signify a decrease in predicted risk of a large outbreak. The color gradient denotes the actual values of the covariates, with red representing high values and blue representing low values.

In both models, *prevalence of open defecation* and *proportion of people using safely managed sanitation services* emerged as the most influential predictors. While the ranking of secondary features varied slightly between models, urban population (%) and time since 2000 consistently ranked within the top five predictors, suggesting strong temporal and demographic drivers of the disease burden. Examples of the model-specific risk factors and protective factors associated with large versus small outbreaks for countries in Oceania are presented in Figures 8 and S6.

**Figure 8:**
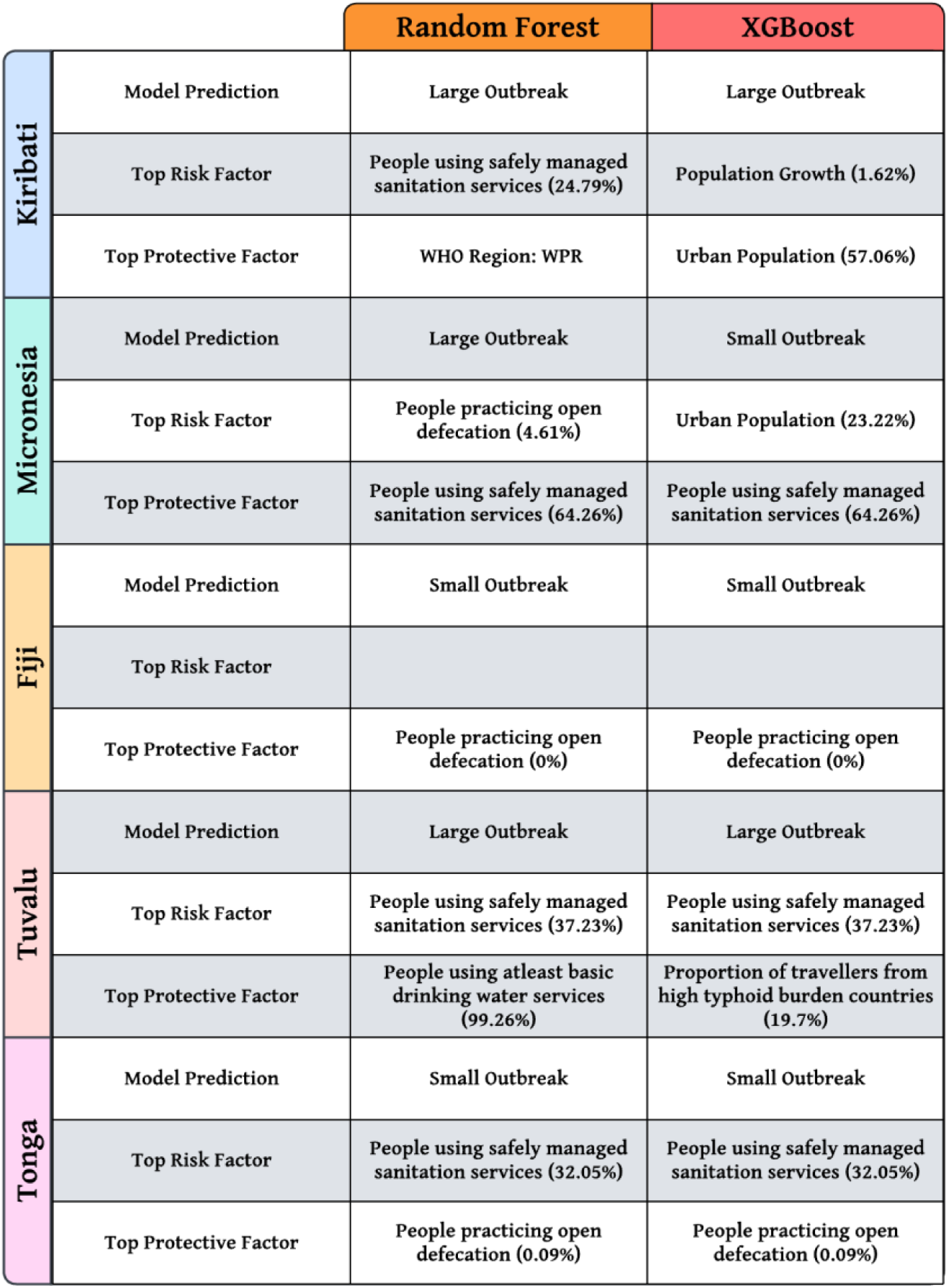
Model-specific influential predictors of typhoid outbreak scale in Oceania identified using SHAP analysis. For each country, Random Forest and XGBoost model predictions (large vs small outbreak) are shown alongside the top risk factor and top protective factor identified through SHapley Additive exPlanations (SHAP) analysis. Percentages indicate the World Bank estimated value of that predictor in 2023.

## Discussion

In this study, we analyzed a comprehensive dataset of recent typhoid fever outbreaks with time series data to identify groups of outbreaks with similar features using unsupervised machine learning methods. We then characterized the size and duration of outbreaks to establish a threshold for identifying the onset of an outbreak and triggering a public health response. To inform preparedness and response planning, we also trained supervised machine learning models to predict whether a typhoid outbreak is likely to be small or large, given the characteristics of the country in which it occurs.

Each of the 34 unique outbreaks in the *Koh et al.* dataset involved at least six total cases or two blood-culture-confirmed cases and lasted for at least 7 days. Based on these shared characteristics, we propose the following operational definition to identify typhoid fever outbreaks: An outbreak is considered to have occurred if six or more typhoid cases (which include suspected, probable, and laboratory-confirmed cases), or at least two blood-culture-confirmed typhoid cases, are reported in a surveillance unit within a 7-day period. The outbreak is considered to have ended if fewer than six typhoid cases (which includes suspected, probable, and laboratory-confirmed cases), and fewer than two blood-culture-confirmed typhoid cases, are reported in a surveillance unit for at least two consecutive weeks. Establishing a threshold could aid in the early detection of typhoid fever outbreaks, thereby facilitating the prompt initiation of public health interventions to mitigate their impact.

Our proposed criteria for determining the start of a typhoid fever outbreak are consistent with the detection thresholds for suspected and confirmed cholera outbreaks. The WHO defines a suspected cholera outbreak as *“two or more suspected cholera cases, or one suspected cholera case with a positive RDT result, reported in a surveillance unit within seven days”*; a probable outbreak as *“the number of suspected cholera cases with a positive rapid diagnostic test (RDT+) achieves or surpasses a defined threshold set by the WHO, taking into account the number of suspected cases tested”*; and a confirmed cholera outbreak as *“a surveillance unit having at least one locally acquired, confirmed cholera case”* [28, 29].

Based on the clustering analysis, we observed that outbreak clusters differed in terms of their scale, enabling a clear distinction between small and large typhoid fever outbreaks on the basis of total cases (<250 or ≥250) alone. Our sensitivity analyses demonstrated that the identification of countries at risk for large-scale outbreaks is largely robust to the choice of outbreak-size threshold. Across all thresholds tested (≥100, ≥250, and ≥500 cases), if an outbreak were to occur, high-income countries were consistently classified at risk of small outbreaks, and countries in sub-Saharan Africa were repeatedly identified as having high risk for large outbreaks. This consistency underscores the reliability of our models in capturing global trends in the magnitude of typhoid fever outbreaks. The classification of South Asian and Oceania countries, however, was more sensitive to the threshold. When a lower threshold of ≥100 cases was applied, fewer South Asian countries were classified as high risk (which seems counterintuitive), and we saw greater model disagreements compared to the primary scenario and the ≥500-case threshold scenario. A plausible explanation for this is that the threshold of ≥100 cases for defining a large outbreak cannot effectively differentiate between moderate- and large-scale outbreaks. If many outbreaks in South Asian and Oceania countries have case counts near or just above this threshold, the models may not be able to differentiate them well, and hence, we end up with increased uncertainty and disagreement among the models. This further reinforces our classification of small versus large outbreaks based on the results of the clustering analysis.

There have been studies that aim to define infectious disease outbreaks, particularly for diseases with robust and long surveillance data. One such study by Porzucek et al. [30] employed a hierarchical Bayesian Poisson regression model to derive a Relative Intensity Score (RISc), which quantifies the deviation of reported dengue cases from an expected baseline. Under this framework, a RISc exceeding 0.5 is indicative of dengue outbreak-like conditions, corresponding to case counts at least 60% above the expected baseline. This Bayesian approach has been successfully applied to dengue fever, for which consistent surveillance data exist across 57 countries spanning 1990 to 2023. Typhoid fever surveillance remains incomplete and inconsistent across many endemic regions, limiting the capacity to establish reliable baseline incidence estimates against which annual deviations could be quantified. Several studies have also attempted to predict the severity of infectious disease outbreaks using machine-learning methods. For instance, Meyer et al. [31] assessed the extent to which climatic, biotic, and demographic variables could predict the severity of chikungunya outbreaks, using the basic reproductive number R_0_ as the metric of outbreak intensity. They defined a chikungunya outbreak as having ≥ 50 cases, fit a mechanistic transmission model to infer R_0_ for each outbreak, and then used supervised machine-learning regression models to evaluate whether pre-outbreak climatic and ecological factors could explain variation in R_0_. Their analyses showed that while climatic, demographic and biotic predictors were associated with R_0_, they had limited ability to predict it. In contrast, our study applies a ≥ 250-case threshold to distinguish small-versus large-scale typhoid fever outbreaks and uses supervised machine-learning classification models to estimate the probability of a large outbreak. While our models exhibited stronger predictive performance, we found that WaSH-related parameters were the dominant contributors to predicting typhoid outbreak scale. Another study by Tadesse et al. [32] used household-level WaSH variables to build a composite variable (“Better” and “Not Better”) through machine learning algorithm and then applied a Cox regression model to evaluate typhoid fever risk based on this composite variable. While a part of their work is focused on deriving an interpretable household-level WaSH rule using a machine learning algorithm to identify higher-risk households, our study applies WASH, demographic, and temporal predictors at the population level to identify regions at greatest risk of large-scale typhoid outbreaks. Both studies use classification-based approaches and optimized thresholds, with Tadesse et al. [32] addressing household-level risk and our work predicting outbreak scale at the national level to inform preparedness efforts.

Our study has several limitations that should be considered while interpreting the results. First, our definition of outbreak onset was based on absolute case counts over a specified time period, without incorporating a population denominator. This was due to inconsistent reporting of population data across the included studies, which limited our ability to express outbreak magnitude in terms of standardized incidence rates (e.g., cases per 100,000 person-years). Researchers and public health authorities should assess what the expected incidence of typhoid fever in a given area is before adopting any specific outbreak threshold. Second, our machine learning models were designed to predict the risk of large outbreaks rather than the probability of an outbreak occurring. Given the limited availability of typhoid surveillance data, particularly for non-outbreak periods, our ability to model the baseline probability of outbreak occurrence is constrained. Third, time series data were not available for outbreaks reported in the Kim et al. and Appiah et al. studies. In these cases, we assumed the outbreaks aligned with our operational definition despite the absence of temporal detail, which may introduce uncertainty. Fourth, we have assumed that the relative volume of air travel between countries is fairly stable over time. While this assumption may be reasonable for most years, it may not hold for certain periods — particularly in 2021 and 2022 — when global air travel was significantly disrupted due to the COVID-19 pandemic. As a result, the air travel data for these years may not accurately reflect actual mobility patterns, potentially affecting the accuracy of outbreak projections for those periods. Although there are some limitations in using air travel data, they are unlikely to substantially affect the results, given the robustness of the model predictions. Fifth, the country-level metrics we used to predict outbreak scale may not be completely representative of sub-national level metrics. Finally, although the supervised machine learning models demonstrated high overall accuracy, their relatively low precision scores during model training suggest a tendency toward falsely large outbreak classifications. Going forward, we could seek to improve the accuracy of our models by incorporating seroincidence and climatic variables and using subnational data when possible.

By analyzing a systematic dataset of typhoid fever outbreaks, we established case thresholds to define the onset and conclusion of outbreaks, which can be used to support their early detection and response. We also used machine learning methods to distinguish between small and large outbreaks and predict this classification on the basis of widely available population metrics. These predictions can be used to identify regions vulnerable to large-scale typhoid fever outbreaks, thereby supporting better preparedness, such as TCV stockpiling, for these events in the future.

## Data Availability

All data underlying the findings of this study are publicly available from previously published sources. The datasets analyzed are from: "Time series data on typhoid fever incidence during outbreaks from 2000 to 2022" (Scientific Data, doi: 10.1038/s41597-024-04289-7 data DOI: 10.17605/OSF.IO/N9CKE), “Typhoid Outbreaks, 1989–2018: Implications for Prevention and Control” (Am J Trop Med Hyg, doi: 10.4269/ajtmh.19-0624 data available in Table 1), and “Spatial and Temporal Patterns of Typhoid and Paratyphoid Fever Outbreaks: A Worldwide Review, 1990–2018” (Clin Infect Dis, doi: 10.1093/cid/ciz705 data available in Annex 2 of the Supplementary Material). The code used for data processing and analysis is publicly available at Zenodo (https://doi.org/10.5281/zenodo.19054251)

## Acknowledgements

We thank Dominic Delport, Nick Scott, and Romesh Abeysuriya from the Burnet Institute for helpful feedback in the preparation of this manuscript.

